# Modelling the impact of travel restrictions on COVID-19 cases in Newfoundland and Labrador

**DOI:** 10.1101/2020.09.02.20186874

**Authors:** Amy Hurford, Proton Rahman, J. Concepción Loredo-Osti

**Affiliations:** Biology Department, Memorial University, St. John’s NL A1B 3X9, Canada; Department of Mathematics and Statistics, Memorial University, St. John’s NL A1B 3X9, Canada; Faculty of Medicine, Memorial University, St. John’s, NL A1C 5B8, Canada

**Keywords:** COVID-19, travel restrictions, Newfoundland and Labrador, importations, epidemic model, branching process

## Abstract

In many jurisdictions, public health authorities have implemented travel restrictions to reduce coronavirus disease 2019 (COVID-19) spread. Policies that restrict travel within countries have been implemented, but the impact of these restrictions is not well known. On May 4^th^, 2020, Newfoundland and Labrador (NL) implemented travel restrictions such that non-residents required exemptions to enter the province. We fit a stochastic epidemic model to data describing the number of active COVID-19 cases in NL from March 14^th^ to June 26^th^. We predicted possible outbreaks over 9 weeks, with and without the travel restrictions, and for contact rates 40% to 70% of pre-pandemic levels. Our results suggest that the travel restrictions reduced the mean number of clinical COVID-19 cases in NL by 92%. Furthermore, without the travel restrictions there is a substantial risk of very large outbreaks. Using epidemic modelling, we show how the NL COVID-19 outbreak could have unfolded had the travel restrictions not been implemented. Both physical distancing and travel restrictions affect the local dynamics of the epidemic. Our modelling shows that the travel restrictions are a plausible reason for the few reported COVID-19 cases in NL after May 4^th^.

## Background

In response to the COVID-19 pandemic, travel restrictions have frequently been implemented (Studdert, Hall, and Mello 2020), yet the efficacy of these restrictions has not been established. Some previous studies consider the impact of international travel restrictions (Chinazzi et al. 2020; Wells et al. 2020; Grépin et al. 2021; Russell et al. 2021), but there is a paucity of studies considering restricted travel within a nation (Grépin et al. 2021) making the implementation of travel restrictions controversial for public health authorities (Studdert, Hall, and Mello 2020). Furthermore, the impact of travel restrictions on reducing COVID-19 spread is interwoven with the impacts of other public health measures. For example, the spread of imported cases depends on compliance with self-isolation directives for travellers, local physical distancing, and mask wearing. Travel restrictions were implemented in Newfoundland and Labrador (NL) on May 4^th^, 2020, such that only NL residents and exempted individuals were permitted to enter the province. We use a mathematical model to consider a “what-if” scenario: specifically, “what if there were no travel-restrictions?”, and in doing so, we quantify the impact that the travel restrictions had on the number of subsequent COVID-19 cases in NL.

Mathematical models appropriate for large populations will poorly predict the epidemic dynamics of smaller populations since chance events may dramatically alter an epidemic trajectory when there are only a few cases to begin with (Keeling and Rohani 2008). As such, it is not clear that results describing the impacts of international travel restrictions will also apply within countries, to smaller regions, and to regions with low infection prevalence. Imported infections due to the arrival of infected travellers will have a disproportionately large effect when the number of local cases is few (Russell et al., 2021). To appropriately characterize the impact of the travel restrictions on the COVID-19 outbreak in NL, we use a stochastic mathematical model appropriate for modelling infection dynamics in small populations (Keeling and Rohani 2008), and where a similar modelling approach has been used in other jurisdictions (Plank et al. 2020; Hellewell et al. 2020). Our analysis quantifies the impact of travel restrictions by considering a higher rate of imported infections when there are no travel restrictions, and we use the model to predict the number of cases that could have occurred in NL in the 9 weeks subsequent to May 4^th^.

## Methods

### Model overview

Our model is based on Plank and colleagues (2020) who use a stochastic branching process to model COVID-19 dynamics in New Zealand. Our model describes the epidemiological dynamics of COVID-19 such that NL residents are either susceptible to, infected with, or recovered from COVID-19. Infected individuals are further divided into symptomatic and asymptomatic infections (infectious, no symptoms for the entire infectious period), and individuals with symptomatic infections may be in either the pre-clinical stage (infectious, prior to the onset of symptoms), or the clinical stage (infectious and symptomatic). The categorization of individuals into these infection classes is consistent with previous work (Hellewell et al. 2020; Davies et al. 2020).

Our model assumes that COVID-19 infections may spread when an infectious person contacts a susceptible person. Contact rates when physical distancing is undertaken in response to the pandemic are expressed in relative terms, as percentages of the contact rate relative to pre-pandemic levels. We assume that the pre-pandemic contact rate was equivalent to a basic reproduction number of R_0_=2.4, where the definition of R_0_ for our model is explained in Table 1. Our model assumes that infected travelers that fail to self-isolate enter the population and may infect susceptible NL residents, and the rate of contact between residents and travellers is assumed to be the same as between residents. For individuals that are infectious (those with asymptomatic, pre-clinical and clinical infections), the probability of infection given a contact depends on the number of days since the date of infection (Ferretti et al. 2020), and infectivity further depends on whether the infection is pre-clinical, clinical or asymptomatic (Davies et al. 2020). Individuals with clinical infections are relatively less infectious because these individuals are symptomatic and are more likely to self-isolate.

**Table 1.** Parameter values.

| Quantity | Description | Source |
| --- | --- | --- |
| Fixed |  |  |
| $N_{pop}=523,000$ | NL population size | Estimated as 519,716 in 2016 (Statistics Canada 2017) |
| $\pi=0.15$ | Proportion of infections that are asymptomatic | Estimated as 17% in Byambasuren et al. (2020). Known to take a wide range of values (Saltelli et al. 2020; Nogrady 2020) |
| $\eta_s = 0.25$ | Proportion reduction in infectivity for asymptomatic infections relative to clinical infections | Davies et al. 2020 |
| $c_{iso} = 0.5$ | Proportion reduction in infectivity for individuals with clinical infections due to self-isolation. | Davies et al. 2020 |
| $R_0 = 2.4$ | The pre-pandemic basic reproduction number. This is the number of secondary infections generated by an individual with a pre-clinical infection over their entire infectivity period, when all individuals in the population are susceptible. For our model, the definition of $R_0$ supposes that the level of infectivity corresponding to a pre- | Assumed |
| | clinical infection is retained for the entire duration of the infectivity period (see equation 1 in the ESM). Note that a change in $R_0$ relative to its pre-pandemic level is equal to the same change in the contact rate if all other contributors to $R_0$ (i.e., the recovery rate and the probability of infection given a contact) are assumed unchanged. | |
| $c_1=30\%$ | Contact rate after March 18<sup>th</sup>, 2020, expressed as a percentage of the pre-pandemic contact rate. | Estimated from NL COVID-19 case data (see Figure 2) |
| $c_2$ in [40%, 70%] | Contact rate after May 4<sup>th</sup>, 2020, expressed as a percentage of the pre-pandemic contact rate. | Range considered |
| Sampled |  |  |
| $W(2.83,5.67)$ | Infectivity, which depends on the number of days since the date of infection (Weibull-distributed). | Ferretti et al. 2020 |
| $s \sim \Gamma(6.1, 1.7)$ | The time from date of infection to self-isolation (gamma-distributed). | Plank et al. 2020. Note that $s \approx T_1 + T_2$, where $T_1$ and $T_2$ appear in Plank et al. 2020. |
| $z_1 \sim \text{POIS}(0.008)$ | The number of imported infected individuals per month that fail to self-isolate when travel restrictions are in place after May 4<sup>th</sup> (Poisson-distributed). The mean value is 0.24 infected travellers per month that fail to self-isolate. | Fit to NL COVID-19 case data when $c_2 \leq 60\%$ (see Figure 2) |
| $z_2 \sim \text{POIS}(0.1)$ | The number of imported infected individuals per month that fail to self-isolate when there are no travel restrictions after May 4<sup>th</sup> (Poisson-distributed). The mean value is 3 infected travellers per month that fail to self-isolate. | The mean importation rate is reduced by 92% when travel restrictions are in place since $z_1 = (1-0.92)z_2$. Therefore, the $z_2$ value is consistent with data reporting a 92.2% decrease in the number of passengers arriving at St. John's airport (NL) in June 2020 relative to June 2019 (Gushue 2020). Equivalently, this assumption can be stated as that without travel restrictions the importation rate is 12.5 times greater ($=0.1/0.008$). |

Similar to models developed by other researchers, our model is formulated as a continuous time branching process (Arino et al. 2020; Hellewell et al. 2020; Plank et al. 2020). A branching process is a type of stochastic model where on any given simulation run, the predicted epidemic may be different since the epidemiological events, and the timing of these events, take values drawn from probability distributions. For example, our model assumes that the number of new infections generated by an infectious person follows a conditional Poisson distribution with a mean that depends on physical distancing, the number of susceptible individuals in the population, the type of infection the infected individual has (asymptomatic, pre-clinical, or clinical), and the number of days since the date of infection (see equation 1 in the Electronic Supplementary Material - ESM). Most other aspects of our model, for example, the timing of new infections, are similarly stochastic, each described by probability distributions that have appropriate characteristics, and are fully described in the ESM. An overview of the model and all parameter values are given in Figure 1 and Table 1.

**Figure 1.**
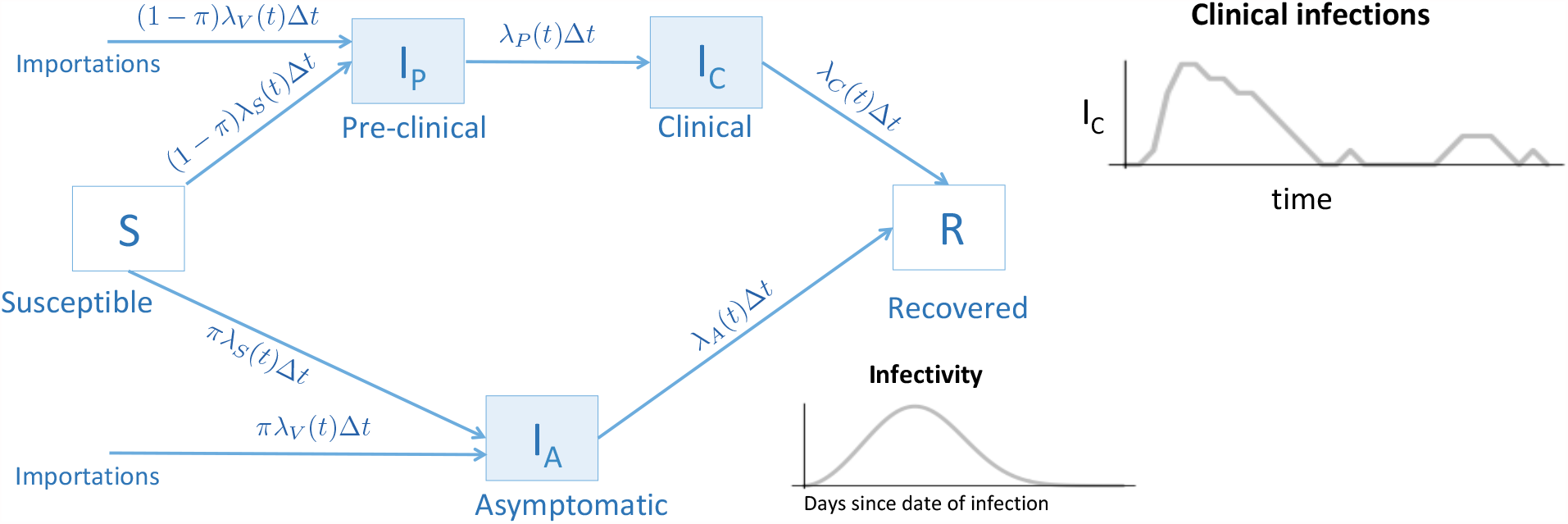
Model diagram. Uninfected individuals (white boxes) are either susceptible to infection, S, or recovered, R. Susceptible individuals become infected at mean rate, λ_S_(t)Δt, where the event that an infection occurs is sampled from a distribution since the model is stochastic. Recovered individuals cannot be re-infected. Infected travellers that fail to self-isolate enter the population at a mean rate, λ_V_(t)Δt. When a new infection occurs, a proportion, π, of these newly infected individuals are asymptomatic, where the number of individuals with asymptomatic infections at any time is I_A_. Alternatively, a proportion, 1-π, of infected individuals will eventually develop clinical symptoms, although these individuals are initially pre-clinical (without symptoms), and the number of individuals that are pre-clinical at any time is I_P_. At a mean rate, λ_P_(t)Δt, individuals with pre-clinical infections develop clinical infections (with symptoms). Individuals with asymptomatic, pre-clinical, and clinical infections are infectious (blue boxes), and infectivity depends on the type of infection, and the number of days since the date of infection. Finally, both individuals with asymptomatic and clinical infections recover at mean rates λ_A_(t)Δt and λ_C_(t)Δt, respectively. See the ESM for further details.

Our model does not consider age-structure or contact rates between individuals in the population that vary in space and time, due to, for example, attending school or work. This latter model limitation is discussed in the *Discussion* section. We intentionally limit the complexity of our model, since when additional parameters are added to a model the uncertainty in the predictions builds up, potentially to the point where the predictions may become useless (Saltelli et al. 2020). The model is implemented in R and the code is publically available at https://doi.org/10.6084/m9.figshare.12906710.v2.

#### Travel restriction scenarios

We assumed that the rate that infected individuals enter NL after May 4^th^, and fail to self-isolate, is Poisson-distributed with a mean, z_1_ = 3 (no travel restrictions) and z_2_ = 0.24 per month (with travel restrictions). The assumed mean rate with travel restrictions yields model predictions compatible with the reported number of cases of COVID-19 in NL after May 4^th^ (see Figure 2). These parameter values, z_1_ and z_2_, imply that with the travel restrictions the number of infected travellers arriving in NL and failing to self-isolate is reduced by 92%; or equivalently, without the travel restrictions the number of infected travellers arriving in NL and failing to self-isolate is 12.5 times greater. The mean rates that infected travellers enter NL and fail to self-isolate (z_1_ and z_2_) are compound parameters consisting of three components: (i) the rate that travellers enter NL; (ii) the proportion of travellers that are infected; and (iii) the proportion of infected travellers that fail to self-isolate. We do not resolve the individual contributions of these three components to z_1_ and z_2_, however we note that only (i), the rate that travellers enter NL, likely changes when travel restrictions are in place. We assumed that infected travellers may be asymptomatic or pre-clinical, as symptomatic travellers are assumed to self-isolate. The proportion of infections that are asymptomatic is assumed to be the same for both travellers and NL residents. The mean rate that infected travellers enter NL is assumed to be constant over time and the origin cities of the travellers is not considered.

**Figure 2.**
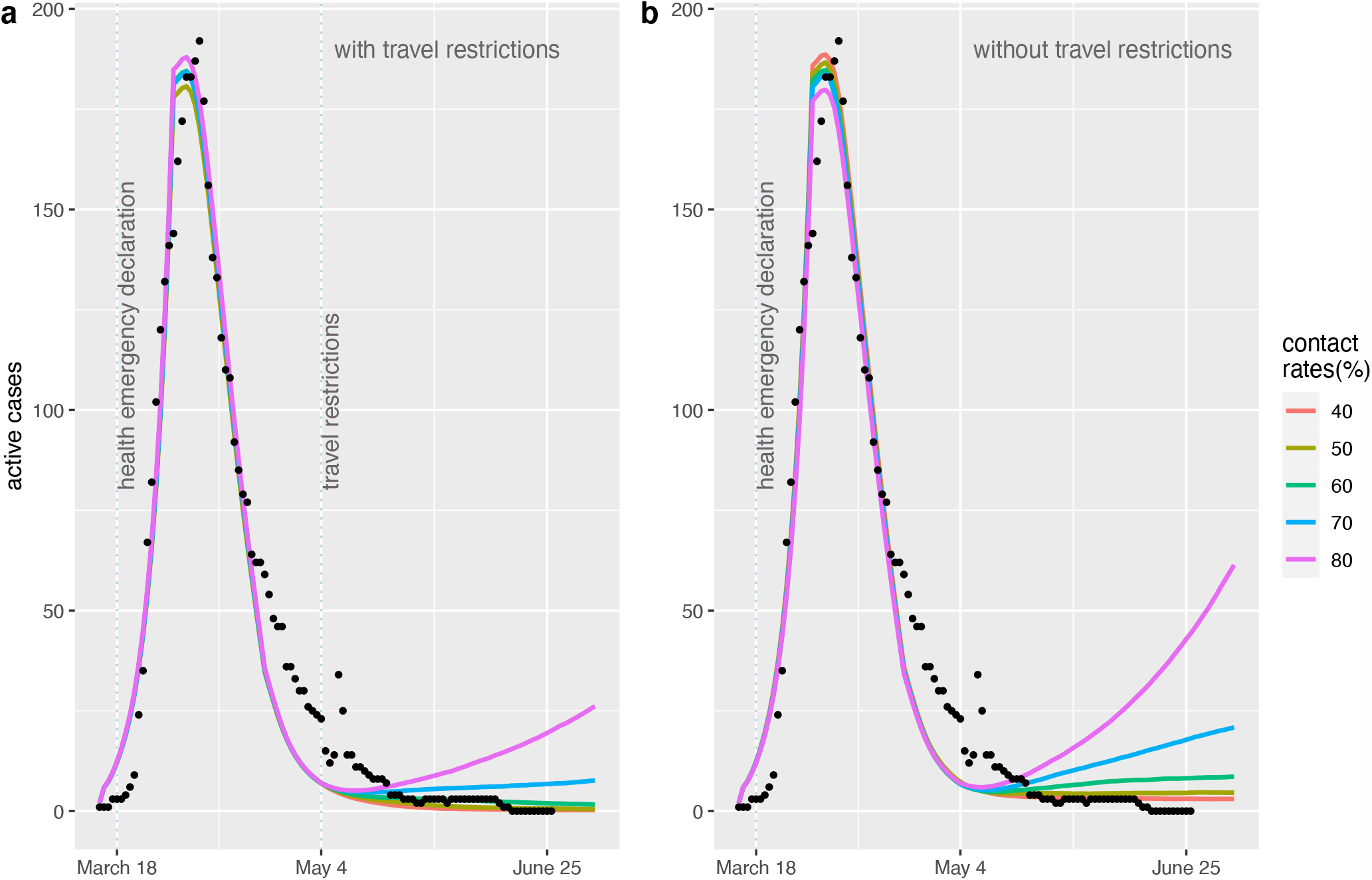
The predicted mean number of active COVID-19 cases (lines) agrees well with the reported numbers of active COVID-19 cases in NL from March 16^th^ to June 26^th^ (dots) prior to the implementation of the travel restrictions on May 4^th^. After May 4^th^, we consider an alternative past scenario where no travel restrictions were implemented (b). Both with (a) and without (b) the travel restrictions, we consider different levels of physical distancing, represented as percentages of the daily contact rate at the pre-pandemic level (coloured lines). Each coloured line is the mean number of active clinical cases each day calculated from 1000 runs of the stochastic model, which considers variability in the timing and changes in the number of individuals with different COVID-19 infection statuses.

### Epidemiological data and public health measures

From March 14^th^ to June 26^th^, 2020, the government of NL reported the number of active COVID-19 cases during media updates and on the Newfoundland and Labrador Pandemic Update Data Hub (for the relevant data, see also Berry et al. 2020). A copy of the data that was used for our analysis is archived with our code (Hurford, Rahman, and Loredo-Osti 2020). In addition to the travel restrictions enacted on May 4^th^, legislation and public health recommendations that would have affected both the importation rate of COVID-19 to NL, and the spread of infections in the community are summarized in Table 2. We assumed that the contact rate between NL residents changed after March 18, 2020, when a public health emergency was declared in NL.

**Table 2.** Public health measures implemented in Newfoundland and Labrador, March 6 - July 3, 2020.

| **Measures affecting the importation rate** |  |
| --- | --- |
| March 20, 2020 | All individuals arriving from outside NL must self-isolate for 14 days (North American COVID-19 Policy Response Monitor: Newfoundland and Labrador, 2020) |
| May 4, 2020 | All individuals are prohibited from entering NL except:<br>Residents of NL<br>Asymptomatic workers and individuals subject to the Exemption Order.<br>Individuals who have been permitted entry to NL, due to extenuating circumstances, approved in advance by the Chief Medical Officer of Health.<br>Individuals arriving from outside NL must self-isolate for 14 days, be available for contact by public health, and complete a travel declaration form at the point of entry. (Special Measures Order - Travel, May 15, 2020). |
| July 3, 2020 | Atlantic bubble: Interprovincial travel without the requirement to self-isolate permitted in Newfoundland and Labrador, New Brunswick, Prince Edward Island and Nova Scotia, for residents of Atlantic Canada (Atlantic Provinces form Travel Bubble, 2020). |
| **Measures affecting community spread** |  |
| March 6, 2020 | Any resident with symptoms asked to stay at home and complete the self assessment tool (North American COVID-19 Policy Response Monitor: Newfoundland and Labrador, 2020). |
| March 18, 2020 | Alert level 5*. State of emergency declared. Residents advised to practice physical distancing and only leave their homes for essential purposes. Only essential businesses open. Gatherings of more than 50 prohibited. Restaurants are takeout only. (Public Health Promotion and Protection Act; *inferred as alert levels not yet defined) |
| March 30, 2020 | Gatherings of more than 5 prohibited (North American COVID-19 Policy Response Monitor: Newfoundland and Labrador, 2020). |
| May 11, 2020 | Alert level 4 (see 'A foundation for living with COVID-19'). Households are permitted to form 'double bubbles'. Gatherings of up to 10 people, reopening of parks and certain businesses. Childcare services operating at 50% (North American COVID-19 Policy Response Monitor: Newfoundland and Labrador, 2020). |
| May 29, 2020 | Six more people can be added to 'double bubbles' (North American COVID-19 Policy Response Monitor: Newfoundland and Labrador, 2020). |
| June 10, 2020 | Alert level 3. Gatherings of up to 20 people, responsible intra-provincial travel, and medium-intensity sports permitted. Childcare services operating at 70% (North American COVID-19 Policy Response Monitor: Newfoundland and Labrador, 2020). |
| June 25, 2020 | Alert level 2. Occupancy and gatherings limited to 50 people, with physical distancing (including funerals, weddings, burials, indoor pools, gyms, movie theatres, bowling alleys, etc). Wakes, karaoke and dance |
|  | floors not allowed. Virtual delivery of health care encouraged. (Public Health Advisory June 24, 2020). |

#### Model calibration

We assumed that prior to March 18, 2020, the pre-pandemic basic reproduction number was R_0_=2.4, where the assumed value of R_0_ affects how quickly the epidemic would grow. All model parameters except the contact rate from March 19^th^ to May 4^th^, c_1_, were estimated independently of the NL COVID-19 case data (see Table 1). The contact rate, c_1,_ is expressed as a percentage relative to the pre-pandemic contact rate (as implied by the pre-pandemic R_0_ assuming all other contributors to R_0_ are fixed). To fit c_1_ given the data, we assumed that all clinical cases were reported, which is a reasonable assumption given the low number of cases reported in NL (for a model that considers unreported cases, see Liu et al. 2020). We estimated c_1_ by observing that c_1_ = 30% resulted in an agreement of the model with the epidemic data (further details of the model calibration are provided in the ESM).

### Output variables

To determine the impact of travel restrictions, we characterize clinical infections occurring in NL after May 4^th^ as:

- Prior: the infected individual is part of an infection chain (i.e., a description of who infected whom) that originates from an NL resident infected prior to May 4^th^.
- Travel: the infected individual was infected prior to travelling to NL.
- Local: the infected individual is an NL resident, who did not travel outside the province, and is part of an infection chain that originates from a traveller to NL.

The number of clinical cases that are ‘travel-related’ is calculated as the sum of infections characterized as ‘travel’ and ‘local’. The predicted number of COVID-19 cases refers only to clinical infections, and does not include asymptomatic infections.

## Results

The predicted number of active clinical COVID-19 cases in NL from March 14^th^ to May 4^th^ (Figure 2, lines) broadly agrees with the data describing the number of active COVID-19 cases in NL over this same period (Figure 2, black dots). From May 4^th^ to June 26, 2020, when the travel restrictions were implemented in NL, the NL COVID-19 case data (Figure 2a, black dots) agrees with the model predictions for physical distancing scenarios corresponding to contact rates ≤ 60% of the pre-pandemic level (Figure 2a; coral – 40%, khaki - 50%, and green – 60% lines).

We estimated that with the travel restrictions in place, from May 4^th^ to June 26^th^, 2020 the mean number of COVID-19 cases is reduced by 92% (Table 3). For the different physical distancing scenarios considered, the mean number of cases over the 9 weeks ranged from 14-48 clinical cases (without the travel restrictions), as compared to 1-4 clinical cases (with the travel restrictions; Table 3 and Figure 3a). These model predictions with the travel restrictions in place are consistent with the COVID-19 data for NL for the 9 weeks following May 4^th^ where during this time 2 new cases of COVID-19 were reported.

**Table 3.** Predicted total number of clinical COVID-19 cases in the 9 weeks subsequent to May 4^th^ with and without the implementation of travel restrictions. The prediction intervals represent the simulated 0.025 and 0.975 quantiles.

| Percentage reduction in the contact rate relative to pre-pandemic levels | Predicted clinical COVID-19 cases over 9 weeks |  |  |  |
| --- | --- | --- | --- | --- |
|  | Travel restrictions | No travel restrictions | Magnitude greater without restrictions | Percentage reduction with restrictions |
| **40%** |  |  |  |  |
| Mean | 1.2 | 13.6 | 11.0 | 91.2% |
| Median | 0 | 12 |  |  |
| 95% prediction intervals | [0,9] | [2,35] |  |  |
| **50%** |  |  |  |  |
| Mean | 1.5 | 18.1 | 12.0 | 91.7% |
| Median | 0 | 15 |  |  |
| 95% prediction intervals | [0,11] | [3,53] |  |  |
| **60%** |  |  |  |  |
| Mean | 2.1 | 27.8 | 13.5 | 92.4% |
| Median | 0 | 23 |  |  |
| 95% prediction intervals | [0,17] | [3,79] |  |  |
| **70%** |  |  |  |  |
| Mean | 3.7 | 47.9 | 13.0 | 92.3% |
| Median | 0 | 35 |  |  |
| 95% prediction intervals | [0,33] | [3,159] |  |  |
|  |  |  | Mean = 12.4 | Mean = 91.9% |

**Figure 3.**
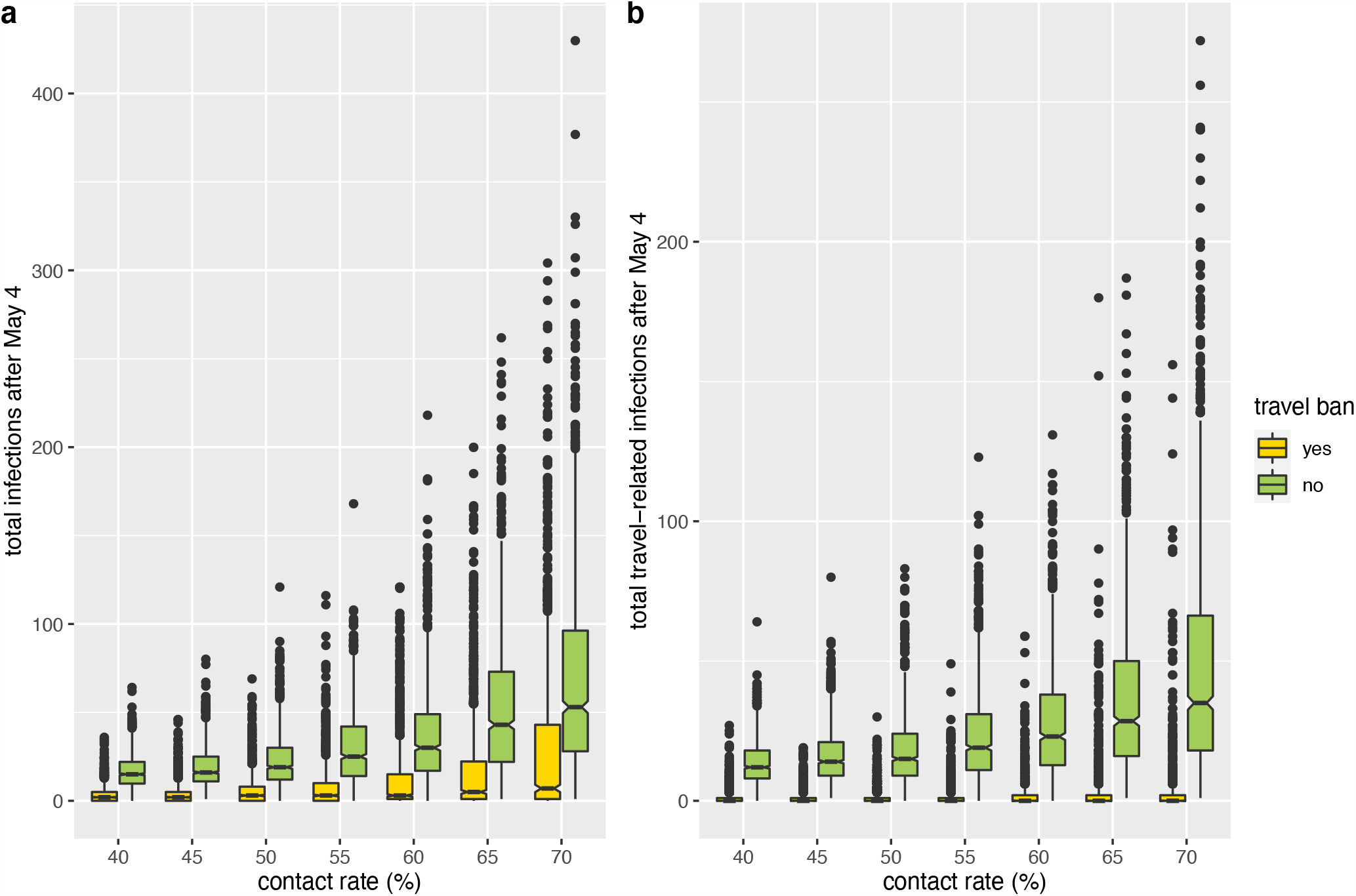
The total predicted number of COVID-19 cases in NL occurring over 9 weeks beginning on May 4^th^ when travel restrictions are implemented (yellow boxes) is much less than the total number of cases occurring over this same period if the travel restrictions were not implemented (green boxes). The total number of COVID-19 cases occurring during the 9 weeks subsequent to May 4^th^ is highly variable, and without the implementation of the travel restrictions there is a higher risk of a large outbreak (also see Table 3 - 95% prediction intervals). When the travel restrictions are implemented, almost all of the cases occurring during the 9 weeks subsequent to May 4^th^ are due to infected individuals present in the community prior to May 4^th^. Travel-related cases are all cases remaining after the ‘prior’ cases are removed (b). The contact rate is expressed as a percentage of the pre-pandemic contact rate. For each simulation, chance events affect the number of individuals that change COVID-19 infection statuses and the timing of these changes. The horizontal lines are medians, the colored boxes are 1.58 times the interquartile range divided by the square root of n, the whiskers are 95% prediction intervals, and the dots are outliers for the n=1000 simulation outcomes.

Without the travel restrictions, the number of clinical cases during the 9 weeks can be very large (Table 3 and Figure 3a). Specifically, for a contact rate at 60% of its pre-pandemic level, the upper limit on the 95% prediction interval for the number of clinical cases over the 9 weeks is 79 (without the travel restrictions) and 17 (with the travel restrictions; Table 3, Figure 3a). The impact of the travel restrictions is even more substantial when only travel-related cases are considered (Figure 3b) since almost all infections arising when the travel restrictions are implemented are attributed to infection chains that arise from an NL resident infected prior to May 4^th^. The mean number of cases of each infection type: ‘prior’, ‘travel’ and ‘local’ are shown in Figure 4.

**Figure 4.**
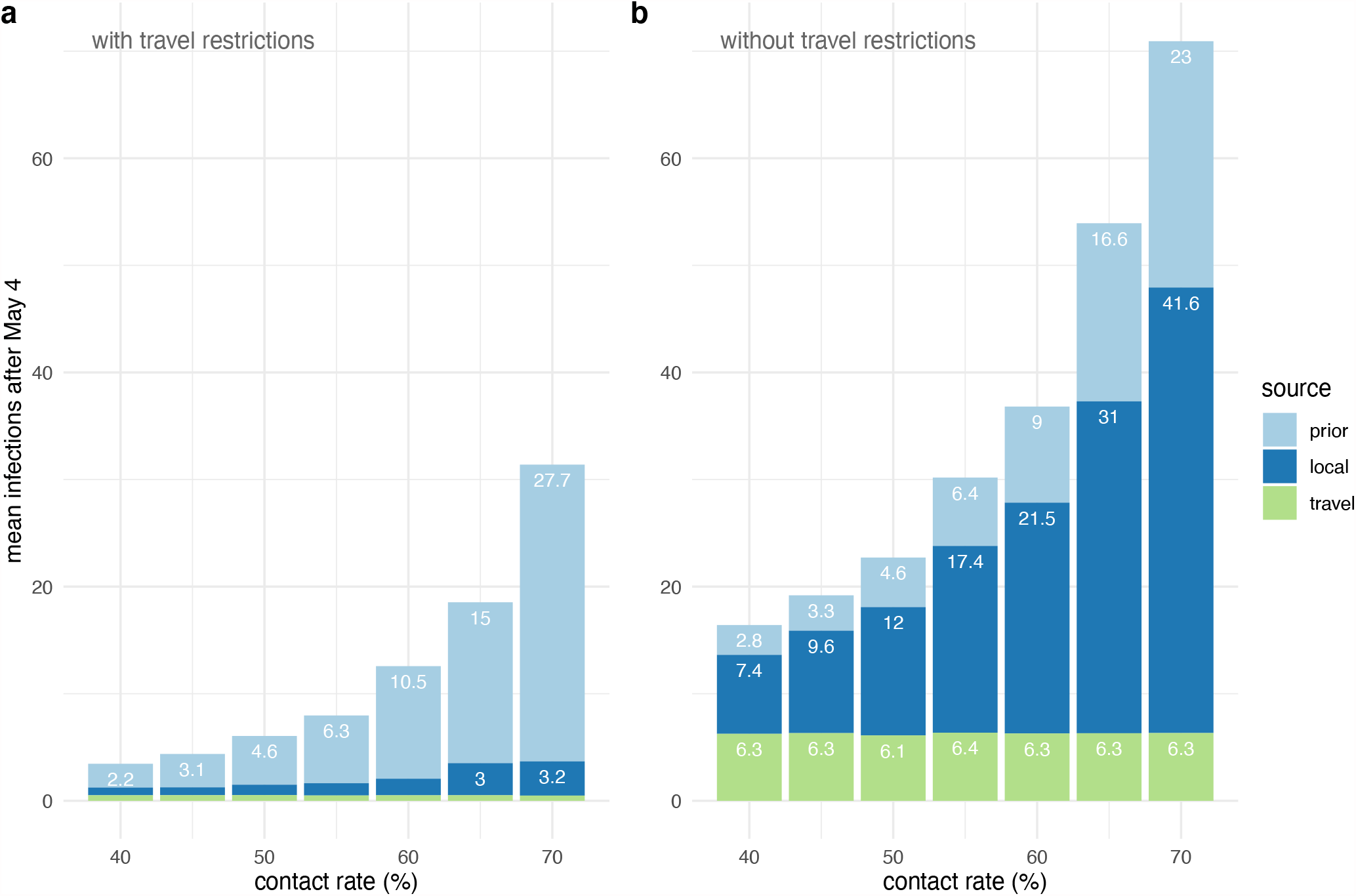
The breakdown into three different sources of COVID-19 cases occurring in NL over 9 weeks. We compare simulation results with travel restrictions (a) and without travel restrictions (b). The source of infections is either: an individual infected prior to May 4^th^ (‘prior’, light blue); an individual that was infected prior to entering NL (‘travel’, green); or a NL resident that did not travel, but is part of an infection chain where the initial infectee is a traveller that entered NL after May 4^th^ (‘local’, dark blue). Our model assumptions are reflected by the difference in the number of COVID-19 cases occurring in travellers over the 9 weeks (green bars): approximately 1.5 with travel restrictions (a), as compared to 6.3 without travel restrictions (b). These infected travellers seed infection chains in the NL community resulting in a larger number of NL residents infected when the travel restrictions are not implemented (dark blue bars). Both with and without the travel restrictions, the number of cases due to prior infection in the NL community is similar (light blue bars). The contact rate is expressed as a percentage of the pre-pandemic contact rate.

## Discussion

Our model predictions broadly agree with the data describing the number of active COVID-19 cases in NL reported from March 14^th^ to May 4^th^, and from May 4^th^ to June 26^th^ if contract rates are 60% or less relative to pre-pandemic levels (Figure 2). Our modelling shows that implementing the travel restrictions on May 4^th^ reduced the number of COVID-19 cases by 92% over the subsequent 9 weeks (Table 3). Furthermore, without the travel restrictions, large outbreaks are much more likely (Table 3 – 95% prediction intervals; Figure 3a). Travel restrictions alone may be insufficient to limit COVID-19 spread since the level of physical distancing undertaken by the local community, which affects the contact rates between residents, is also a strong determinant of the outbreak size (Figures 2-4).

We found that the decrease in the mean number of clinical infections when the travel restrictions were enacted (a 92% reduction; see Table 3) was nearly exactly equal to the reduction in travel due to the travel restrictions (a 92% reduction; see Table 1). This equivalency was expected due to the hypothesized linear relationship between the importation rate and the mean outbreak size as noted in Anderson et al. 2020. A consequence of this linear relationship is that any relative changes in the mean outbreak size are expected to be equal to the relative changes in the importation rate (with travel restrictions relative to without restrictions and visa versa). The assumptions and characteristics of our model that give rise to this linear relationship are discussed in Table 4 along with examples of conditions where these assumptions would be violated.

**Table 4.** A list of the assumptions and characteristics of our model that give rise to the linear relationship between the importation rate and the mean outbreak size. The linear relationship is that I_tot_ =λ_v_I_1_, where I_tot_ is the mean total number of cases, λ_v_ is the importation rate, and I_1_ is the mean number of cases that arise from one importation.

| Model assumption | Example where the model assumption is violated | Effect of violating the assumption on outbreak size |
| --- | --- | --- |
| Mixing between individuals in the population is homogeneous. | A group of travellers, all of whom are infected, fail to self-isolate, but also travel everywhere together and | No matter what the size of the group, the resulting outbreak will be of similar size since the contacts of group members are redundant. Here, the |
| Homogeneous mixing means that an infected person is equally likely to contact every susceptible person in the population. | contact all of the same people.<br>Mixing is non-homogeneous because group members are constrained to have contacts only amongst the same individuals as the other group members, and not all individuals in the population. | mean outbreak size is not linearly related to the importation rate because a larger group would correspond to a larger number of importations, yet the resulting outbreak would not be much larger. |
| The number of susceptible people is relatively unchanged during the timeframe of interest. | The susceptible population is small, or infection control measures are few. | Infected individuals that arrive later will generate smaller infection chains due to fewer susceptible people to infect. Therefore, the total outbreak size cannot be calculated by summing the size of the outbreaks per importation, since the timing of the importation affects the outbreak size due to that importation. |
| The number of people an infected person contacts is unchanged during the timeframe of interest. | Waning compliance with public health measures; school re-openings. | As above, outbreak sizes per importation cannot be added to determine the total outbreak size because the timing of the importations affects the value of the outbreak size per importation. |
| Infectivity does not change over time. | Seasonality | See above. |
| **Model characteristic** | **A different characteristic** | **Effect of considering the different characteristic** |
| Few 'prior' cases: cases that are not attributable to importations (see *Methods – Output variables*). | High infection prevalence in absence of importations. | The relationship between travel-related cases and the importation rate will be linear, but total infections is the sum of prior cases and travel-related cases, such that the linear relationship will not hold. |
| The quantity of interest is the mean outbreak size. | The quantity of interest is the median or a different quantile. | The linear relationship with the importation rate applies only to the mean outbreak size. As can be observed in Table 2, the linear relationship does not apply to the median or 95% prediction intervals. |

Related research, using North American airline passenger data from January 1, 2019 to March 31, 2020, in combination with epidemic modelling, found that depending on the type of travel restrictions, the effective reproduction number, and the percentage of travellers quarantined, it would take between 37 and 128 days for 0.1% of the NL population to have been infected (Table 2 in Linka et al. 2020). These predicted epidemic trajectories are consistent with our results. However, unlike Linka et al. 2020, we have modelled importations and the NL epidemic dynamics as a stochastic process due to the low infection prevalence in NL at the time of our study.

### Future directions

Our model does not consider spatial structure such that individuals contact each other in schools, workplaces, or ‘bubbles’. The absence of spatial structure in our model may over-estimate the probability of an epidemic establishing and the total number of cases until the outbreak subsides (Keeling 1999). Related research, however, does consider spatially structured interactions in workplaces, businesses and schools, and concludes that without the travel restrictions implemented in NL on May 4^th^ the number COVID-19 cases would have been 10 times greater (Aleman et al., 2021) which is in close agreement with our results that the number of cases would have been 12.5 times greater (Table 3): a result that arises due to our parameterization of the importation rate without travel restrictions as 12.5 times greater than with travel restrictions (Tables 1 and 4). Travel restrictions are one of several approaches available to health authorities for COVID-19 management. Future research should consider the role of travel restrictions, testing, contact tracing and physical distancing, as elements of comprehensive approach to the best management of COVID-19.

### Limitations

We were not able to estimate the rate that infected travellers enter NL, however other research modelling infection dynamics in the origin cities of air travellers to NL found that without travel restrictions a new COVID-19 case would enter NL every other day (Linka et al. 2020). Similarly, we were not able to estimate the percentage of travellers to NL that comply with self-isolation directives. Smith et al. (2020) found that 75% of survey participants reporting COVID-19 symptoms (high temperature and/or cough) also report having left their house in the last 24 hours, violating the lockdown measures in place in the UK at the time, and so non-compliance rates may be quite high. Our analysis does not consider hospitalizations or deaths, however, we note that as of May 4^th^, 2020, NL had experienced 259 clinical cases and 3 deaths. With the contact rate at 80% of its pre-pandemic level and no travel restrictions, we estimate that it would take, on average, 10.2 weeks for a further 259 clinical cases to occur, and although there is evidence that case fatality rates have changed over time (Ledford 2020), it is reasonable to expect a further 3 deaths under these conditions. In contrast, with the travel restrictions in place, it would take more than 6 months (28.1 weeks) for this same number of cases and deaths to accumulate. Thus, with the first COVID-19 vaccines available to the public a year after the beginning of the pandemic, the value of enacting travel restrictions to delay the local outbreak by 6 months is potentially substantial.

## Conclusion

At the time of the implementation of the travel restrictions, there were few COVID-19 infections in NL. Without the travel restrictions, most of the subsequent COVID-19 infections would have been initiated by infected travellers who failed to comply with self-isolation requirements and only the actions of NL residents (i.e., physical distancing), and local health authorities (i.e., testing and contact tracing) would be sufficient to slow the exponential growth of these infection chains in the local community.

## Data Availability

All data and code are archived on FigShare.

https://doi.org/10.6084/m9.figshare.12906710.v2

## Electronic Supplementary Material

### 1. Model description

1. Infected individuals either: (i) will show clinical symptoms at some point during their infection (with probability 1-π), or (ii) will be asymptomatic (with probability π).
2. Individuals that are pre-clinical, clinical, or asymptomatic (see Figure 1) are all infectious with different levels of infectivity given contact with a susceptible person. The infectivity of infected individuals changes depending on the number of days since infection onset and follows a Weibull distribution that is parameterized such that peak infectivity occurs approximately 5 days after the initial infection, and 90% of infections occur between 2.0 and 8.4 days after the infection onset. It is also assumed that 21 days after infection onset an individual is no longer infective. See Ferretti et al. 2020 for a justification of this assumption.
3. Individuals with asymptomatic infections are less likely than pre-clinically infected individuals to infect a susceptible person given a contact, where η_S_ is a coefficient that scales the infectivity of asymptomatic individuals relative to pre-clinically infected individuals.
4. Clinically infected individuals are assumed to self-isolate, which reduces their infectivity by a factor c_iso_ relative to individuals with pre-clinical infections.
5. Infected individuals that will progress to have a clinical infection have an initial period when they are pre-clinical, T_1_. This distribution is the same as the distribution for the period from the date of infection to self-isolation, and is gamma-distributed, s _∼_ Γ(6.1,1.7). Note that we let s≈ T_1_ + T_2_, where T_1_ and T_2_ appear in Plank et al. 2020.
6. Each infected individual *j*, per unit time, generates a Poisson-distributed number of new infections with a mean equal to λ_j_(t) Δt. This mean number of secondary infections depends on the fraction of susceptible people in the population, 1 – N(t)/N_pop_, the type of infection the infective person has, F_j_(t), the infectivity of the infected individual a given number of days since the date of infection, whether the infected person is in self-isolation, and the rate of contacts between individuals in the population. The rate of infection for the j^th^ individual (infected at the time t_j_) on the time interval (t, t+Δt] is λ_j_(t)Δt, where

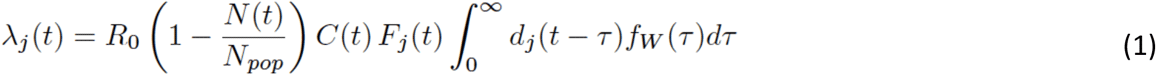

and,

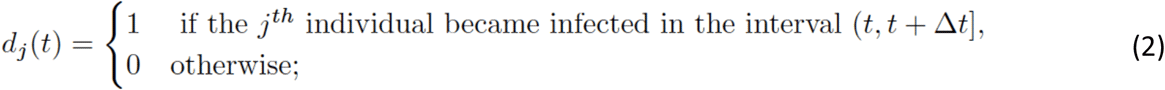

f_W_(τ) is the density of the serial-interval time, W, and F_j_(t) and C(t) are given by,

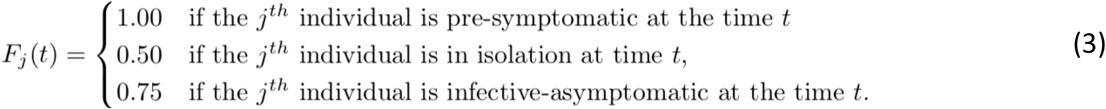

and, C(t), the function that accounts for public health measures is defined as,

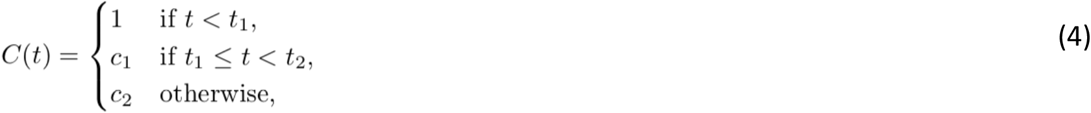

where t_1_ = March 18, 2020 is the date of the declaration of the health emergency in NL, and t_2_= May 4, 2020, the date when we consider scenarios representing different contact rates between NL residents. We performed additional simulations where the number of new infections followed a negative binomial distribution. Our results were strongly consistent with the simulations when the Poisson distribution was assumed (Figure A.3).
7. The time between an individual becoming infected and infecting another individual, the generation time, follows a Weibull distribution with a shape parameter equal to 2.83 and a scale parameter equal to 5.67 (mean value is 5 days). The infection times of all N_j_ secondary infections from an individual j are independent identically distributed random variables from this distribution.
8. On an interval of length Δt, the rate that infected travellers arrive and fail to self-isolate is λ_V_(t)Δt, which follows a Poisson distribution with the parameter λ_V_(t) given as,

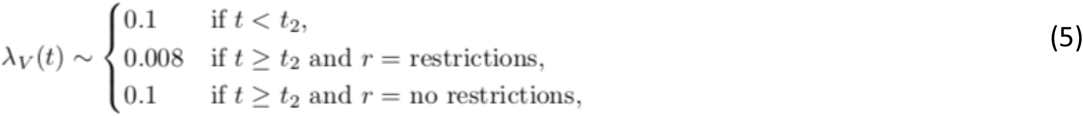

where r = restrictions corresponds to travel restrictions, r = no restrictions corresponds to no travel restrictions, and t_2_ corresponds to May 4, 2020.

The model is a stochastic birth-death process where births correspond to new infections and deaths correspond to the recovery of infected individuals. The counts arise from a non-homogeneous Poisson process, and the model describes a lagged process owing to the consideration of the serial interval distribution. The model is implemented in R using Euler’s method (Gardner 2009).

### Definitions of the mean rates appearing in Figure 1

Susceptible individuals become infected at a mean rate, λ_S_(t)Δt, with λ_S_(t) = Σ_j_ λ_j_(t) where λ_j_(t) is given by equation 1. Infected travellers that fail to self-isolate enter the population at a rate λ_V_(t) (equation 5). At a rate, λ_P_(t)Δt, with λ_P_(t) = Σ_j_ γ_j_^P^(t) individuals with pre-clinical infections develop clinical infections. Finally, both individuals with asymptomatic and clinical infections recover at rates λ_A_(t)Δt with λ_A_(t) = Σ_j_ γ_j_^A^(t) and λ_C_(t)Δt with λ_C_(t) = Σ_j_ γ_j_^C^(t), respectively. The probability of removing the j^th^ individual from the K class in the time interval (t,t+Δt], given that this individual has not been removed before is,

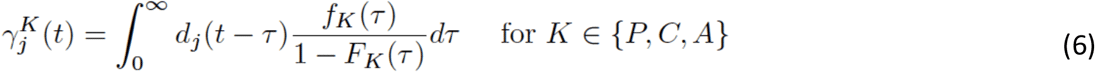

where f_K_(τ) and F_K_(τ) are the density and distribution functions for the time to removal from the K class

### 2. Negative binomial distribution of secondary infections

**Figure A.1.**
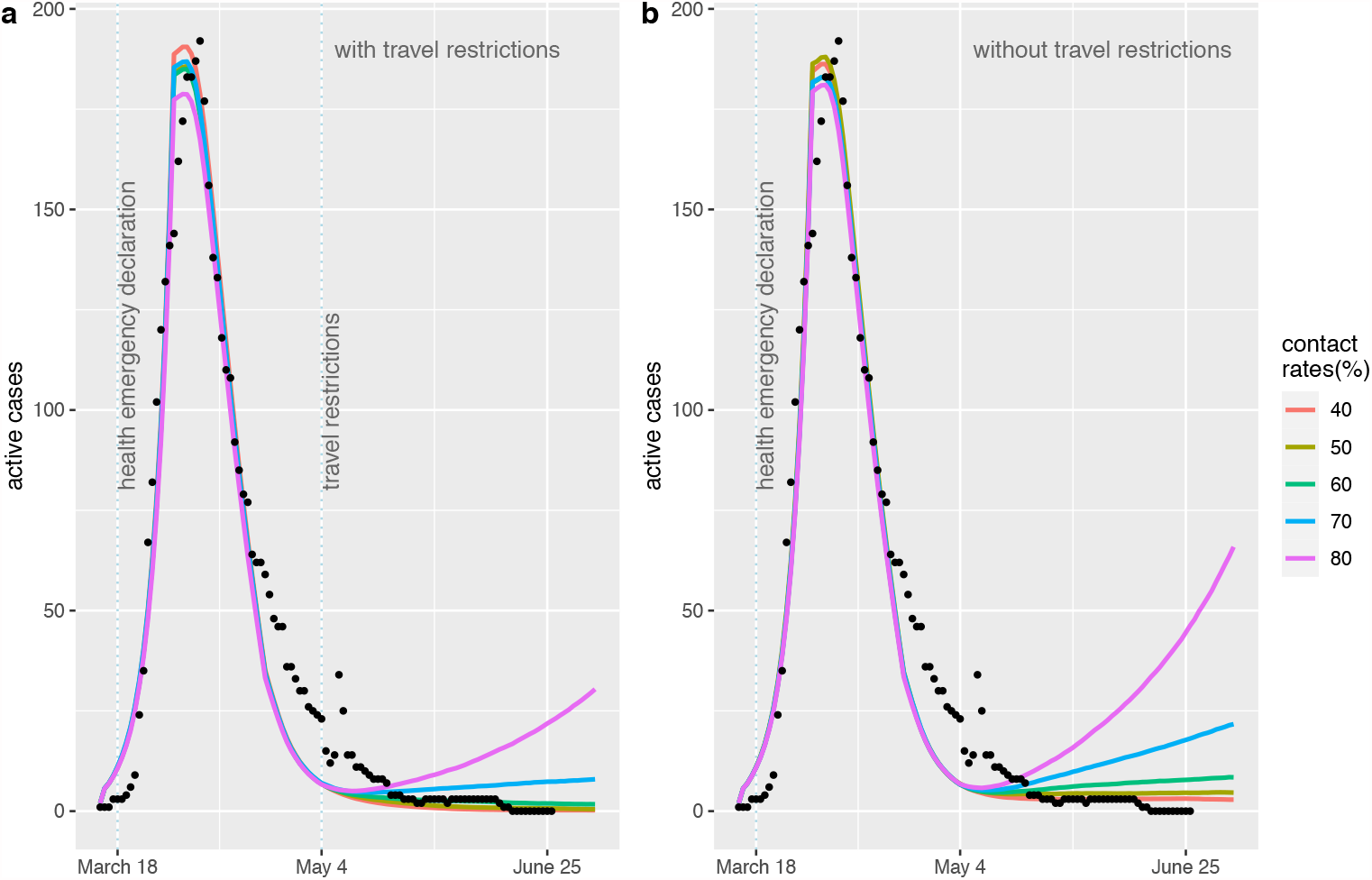
We repeated our simulations assuming that the number of secondary infections followed a negative binomial distribution with k = 0.1 (Endo et al. 2020) rather than a Poisson distribution (see 6. of Model description in this Appendix). For the negative binomial distribution, we set R_0_ = 4.67 so that the model predictions were consistent with the NL data from March 16^th^-June 26^th^, 2020, as shown in this figure.

**Figure A.2.**
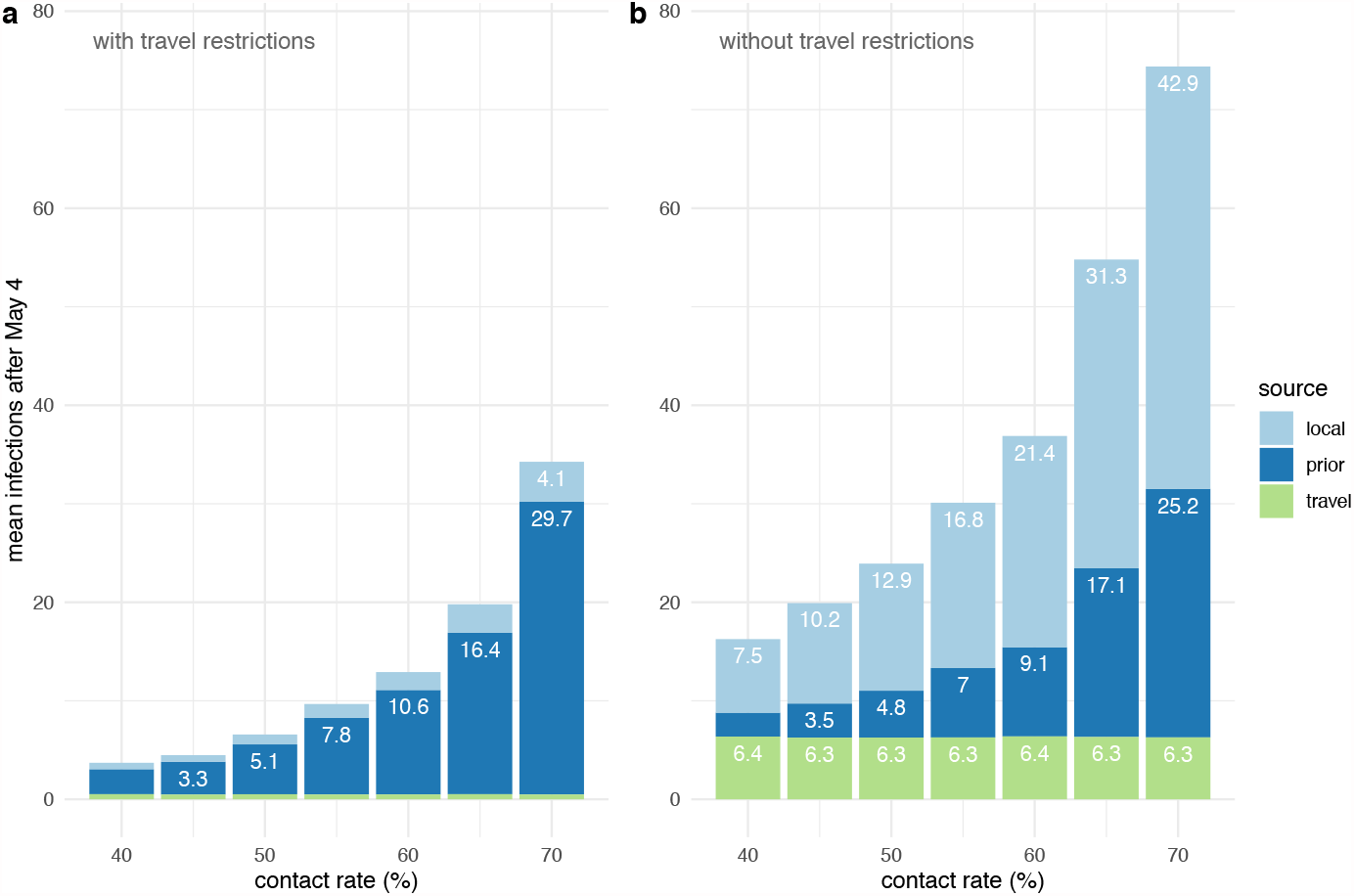
We repeated our simulations assuming that the number of secondary infections followed a negative binomial distribution with k = 0.1 (Endo et al. 2020) rather than a Poisson distribution (see 6. of Model description in this Appendix). For the negative binomial distribution, we set R_0_ = 4.67 so that the model predictions were consistent with the NL data from March 16^th^-June 26^th^, 2020 (Figure A.1). This figure is comparable to Figure 3, which assumed a Poisson distribution of secondary infections.

### 3. Model Calibration

We estimated the percentage of contacts between March 19 and May 4, 2020, relative to the pre-pandemic level as c_1_ = 30%. To estimate c_1_, we used model calibration, where different values of c_1_ were considered and the resulting agreement with the data was observed. In Figure A.3, we show that when c_1_ = 20% (red) the peak number of active cases occurs too early, and the number of active cases during the decline is under-predicted. When c_1_ = 40% (Figure A.3, blue), the number of active cases before and after the peak is over-estimated. For our analysis, we used c_1_ = 30% (Figure A.3, green), as this value was consistent with the epidemic data (black dots). We did not consider a formal fitting algorithm due to the long computational times associated with fitting stochastic models, because we cannot precisely estimate the other model parameters, and because Figure A.3 demonstrates that the estimated c_1_ value is likely between 20 and 40%.

**Figure A.3.**
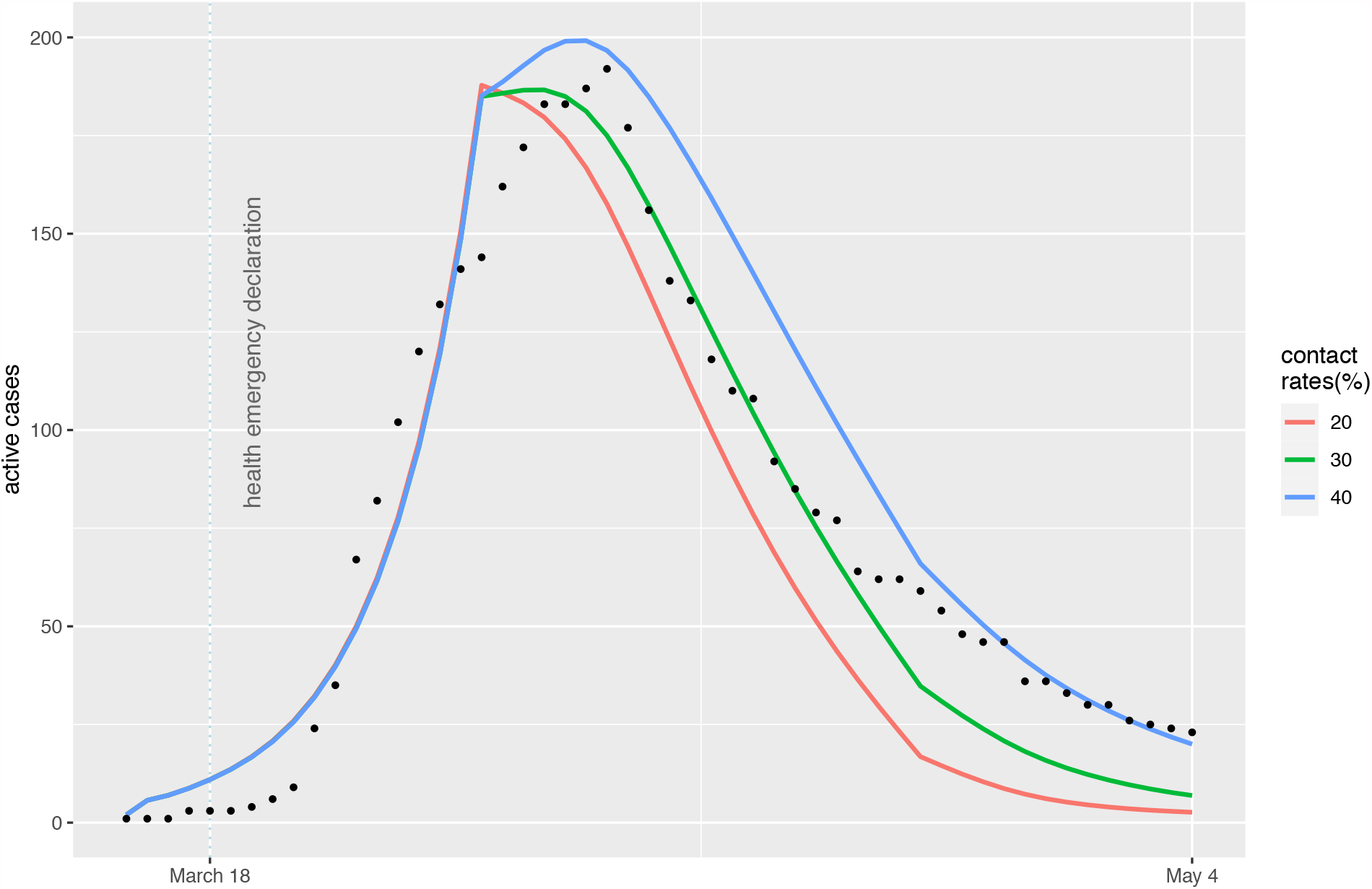
Our analysis assumed the percentage of contacts between March 19 and May 4, 2020 relative to the pre-pandemic baseline was c_1_ = 30% (green line). This value was estimated using model calibration and observing the agreement of the model with the epidemic data (black dots). If c_1_ = 20% (red), the peak number of active cases occurs too early, and the number of active cases during the decline is under-predicted. If c_1_ = 40% (blue), the number of active cases before and after the peak is over-estimated.

